# Unravelling the early warning capability of wastewater surveillance for COVID-19: A temporal study on SARS-CoV-2 RNA detection and need for the escalation

**DOI:** 10.1101/2020.12.22.20248744

**Authors:** Manish Kumar, Madhvi Joshi, Arbind Kumar Patel, Chaitanya G Joshi

## Abstract

Wastewater-based Epidemiological (WBE) surveillance offers a promising approach to assess the pandemic situation covering pre-symptomatic and asymptomatic cases in highly populated area under limited clinical tests. In the present study, we analysed SARS-CoV-2 RNA in the influent wastewater samples (*n* = 43) from four wastewater treatment plants (WWTPs) in Gandhinagar, India, during August 7^th^ to September 30^th^, 2020. A total of 40 samples out of 43 were found positive i.e. having at least two genes of SARS-CoV-2. The average Ct values for S, N, and ORF 1ab genes were 32.66, 33.03, and 33.95, respectively. Monthly variation depicted a substantial rise in the average copies of N (∼120%) and ORF 1ab (∼38%) genes in the month of September as compared to August, while S-gene copies declined by 58% in September 2020. The SARS-CoV-2 genome concentration was higher in the month of September (∼924.5 copies/ L) than August (∼897.5 copies/ L), corresponding to a ∼ 2.2-fold rise in the number of confirmed cases during the study period. Further, the percentage change in genome concentration level on a particular date was found in the lead of 1-2 weeks of time with respect to the official confirmed cases registered based on clinical tests on a temporal scale. The results profoundly unravel the potential of WBE surveillance to predict the fluctuation of COVID-19 cases to provide an early warning. Our study explicitly suggests that it is the need of hour that the wastewater surveillance must be included as an integral part of COVID-19 pandemic monitoring which can not only help the water authorities to identify the hotspots within a city but can provide up to 2 weeks of time lead for better tuning the management interventions.

**HIGHLIGHTS:**

□ Study unravels the early warning potential of wastewater based surveillance of COVID-19.
□ Adequate SARS-CoV-2 RNA were detected despite of limited reported case in the vicinity.
□ Up to 2 weeks of lead is possible from a regular wastewater based COVID-19 surveillance.
□ SARS-CoV-2 RNA was higher in September than August in response to a ∼ 2.2-fold rise in COVID-19 active cases.

**Graphical Abstract:** 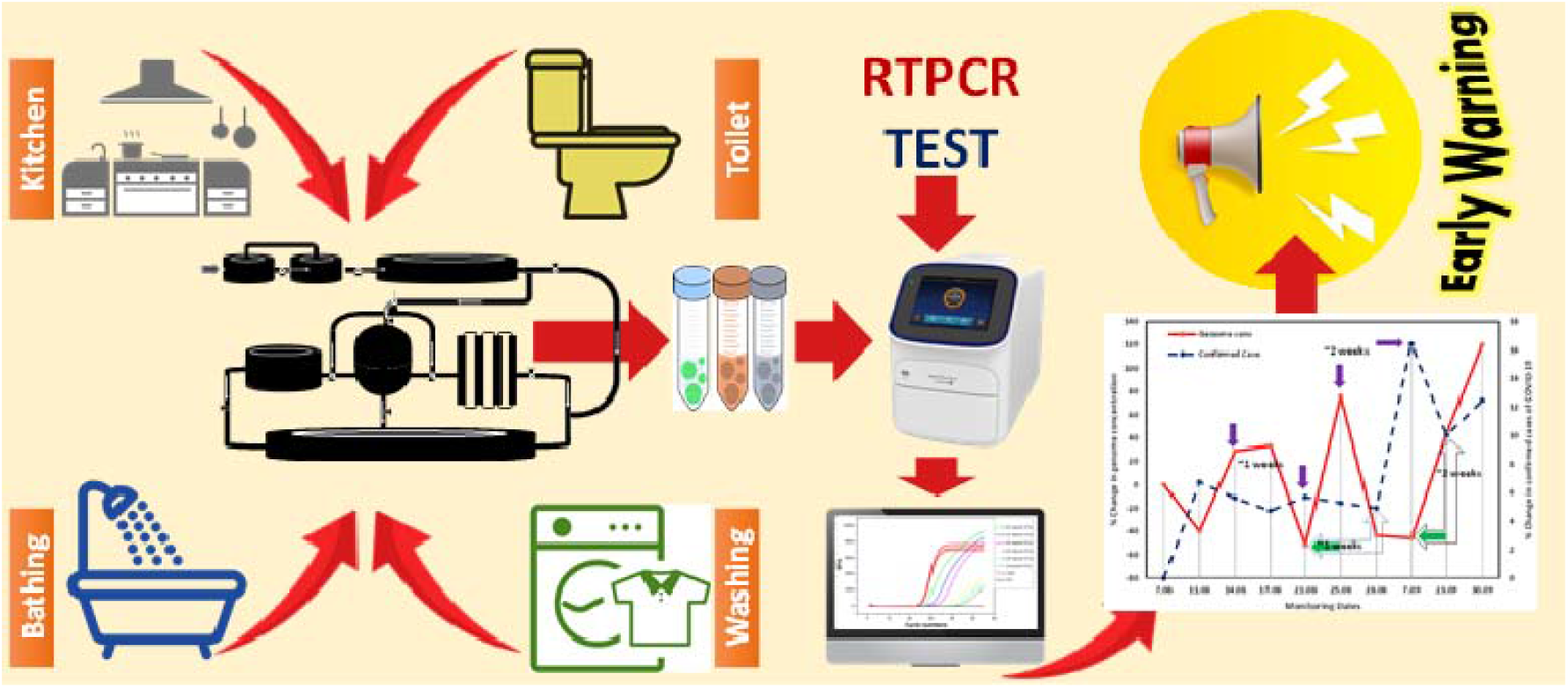

## 1. Introduction

The global pandemic caused by severe acute respiratory syndrome 2 (SARS-CoV-2) disease has led to more than 40 million confirmed cases and >1 million deaths worldwide, covering 216 countries, as of December 10^th^, 2020 (WHO, 2020). The high prevalence of asymptomatic infectious persons is a matter of concern that raises doubt on the available data of active cases based on a clinical survey (Rimoldi et al., 2020; Medema et al., 2020). Therefore, alternative approaches such as wastewater-based epidemiology (WBE) are gaining recognition, and surveillance of SARS-CoV-2 transmission and realtime trend monitoring is being endorsed to trigger pandemic responses by scientific communities (Medema et al., 2020; Randazzo et al., 2020). The SARS-CoV-2 virus replicates in epithelial cells of alveoli and enterocytes of the intestinal lining in human beings due to the expression of ACE2 receptor resulting in respiratory illness and gastrointestinal symptoms such as vomiting and diarrhoea (Ni et al., 2020; Kumar et al., 2020; Gupta et al., 2020; Zhang et al., 2020, Xiao et al., 2020). The clinical symptoms of SARS-CoV-2 infection include cough, breathing problems, diarrhoea, and fever. Different studies suggest that 48 to 67% of deceased persons exhibited SARS-CoV-2 RNA in the stool (Chan et al., 2020; Cheung et al., 2020; Parasa et al., 2020; Wong et al., 2020).

Due to the presence and extended excretion of SARS-CoV-2 RNA in the faecal matter of pre-symptomatic and deceased persons, WBE is gaining attention worldwide to monitor COVID-19, particularly in the developing economies with poor health infrastructure. An earlier investigation on COVID-19 patients revealed the prevalence of SARS-CoV-2 RNA in the stool of a larger population (48.1 %) than patients with gastro-intestinal symptoms (17%) (Cheung et al., 2020). The latter study suggested that asymptomatic persons together with symptomatic persons, discharge viral particles in the excreta finding their way to sewage treatment plants. Interestingly, 18-45% of patients lack symptoms in the case of COVID-19 infection but are capable of transmitting the disease and can adversely affect the actual containment of COVID-19 (Lavezzo et al., 2020; Yang et al., 2020; Mizumoto et al., 2020; Nishiura et al., 2020). Haver and co-workers (2020) reported 6 to 24 times higher infection among asymptomatic and mild symptomatic individuals than confirmed cases at ten different sites in the United States based on surveillance of antibodies to SARS-CoV-2.

The wastewater encompasses SARS-CoV-2 RNA from both asymptomatic and symptomatic patients; therefore, WBE may prove its worthiness for COVID-19 surveillance to forecast the overall pandemic situation. WBE may help in identifying the hotspots and tuning the public health initiatives that will give preparatory time to the regulatory bodies to handle the adverse situation. Further, WBE could provide an early warning of possible re-outbreaks and seasonal outbreaks in the future. The occurrence of SARS-CoV-2 RNA in wastewater has widely been reported from all corners of the world, including Spain, France, Italy, China, Netherlands, Australia, India, and Japan (Randazzo et al., a,b; Wurtzer et al., 2020; Zhang et al., 2020; Medema et al., 2020; La Rosa et al., 2020; Ahmed et al., 2020; Kumar et al., 2020, 2021; Honda et al., 2020). Though the sensitivity of WBE is comparatively less than clinical trials and largely depends on the viral load in the patient’s faecal matter. Though, earlier clues and wide acceptability of WBE suggest that this approach could be superior to clinical surveillance for the early prediction of COVID-19 status for highly populated places. Some earlier studies showed the presence of SARS-CoV-2 in wastewater prior to the first documented report in the study area (Medema et al., 2020; Randazzo et al., 2020; La Rosa et al., 2020). However, to evaluate WBE’s potential as an early prediction tool for COVID-19 pandemic, it is essential to explore the correlation between the SARS-CoV-2 genetic load in wastewater and the number of cases at the district level in each country.

In view of this, the objective of this study was to verify the WBE approach for COVID-19 by comparing the detected concentration of SARS-CoV-2 in wastewater with the COVID-19 cases reported by the clinical surveillance. The detected concentrations of SARS-CoV-2 RNA in wastewater would reflect the true prevalence of COVID-19 infection in the sewer catchment, including clinically undiagnosed patients, while the number of clinically reported cases covers only diagnosed patients and also depends on the number of PCR diagnosis. In the present study, we analysed SARS-CoV-2 RNA in the influent wastewater samples (*n* = 43) from four wastewater treatment plants (WWTPs) in Gandhinagar, India, from August 7^th^ to September 30^th^, 2020, with the following objectives: a) detection and quantification of SARS-CoV-2-RNA in the influent wastewater samples of Gandhinagar to understand the pandemic situation over two months b) biweekly and weekly resolution of the data for two months in genetic material loadings; and c) explicating the potential of WBE for COVID-19 surveillance as a potential tool for identifying hotspots and public health monitoring at the community level.

## 2. Material and Methods

### 2.1 Sampling approach

Influent wastewater samples were collected from four different treatment plants *viz*. Basan, Jaspur, and Sargasan wards, and an academic institution present in the municipal territory of Gandhinagar. The capacity of treatment plants was 2, 10, 10, and 2.36 MLD, respectively. The details of the WWTPs, including their geospatial position, capacity, treatment process, etc., are shown in **Table S1**. The influent wastewater samples were collected from each WWTP first biweekly, followed by weekly for two months, from August to September 2020. A total of forty-three influent wastewater samples collected from four different treatment plants were analyzed for two months. All the samples were collected by grab hand sampling using 250 ml sterile bottles (Tarsons, PP Autoclavable, Wide Mouth Bottle, Cat No. 582240, India). Simultaneously, blanks in the same type of bottle were examined to know any contamination during the transport. *In-situ* water quality parameters (pH, Electrical Conductivity, Dissolved Oxygen, Oxidation-Reduction Potential, and Total Dissolved Solids, Salinity) were examined prior to the sample collection using YSI Multiparameter probe and summarized in **Table S2**. The samples were kept cool in an ice-box until analysis.

### 2.2 SARS-CoV-2 gene detection

#### 2.2.1 Precipitation of viral particle

30 mL samples were centrifuged at 4000×g (Model: Sorvall ST 40R, Thermo Scientific) for 40 minutes in a 50 mL falcon tube followed by filtration of supernatant using 0.22-micron syringe filter (Mixed cellulose esters syringe filter, Himedia). After filtrating 25 mL of the supernatant, it was treated with PEG 9000 (80 g/L), and NaCl (17.5 g/L) was mixed in 25 ml filtrate, and this was incubated at 17°C, 100 rpm overnight (Model: Incu-Shaker™ 10LR, Benchmark). Next day, the mixture was centrifuged at 14000×g (Model: Kubota 6500, Kubota Corporation) for about 90 minutes. The supernatant was discarded after centrifugation, and the pellet was resuspended in 300µL RNase free water. The concentrated sample was kept in 1.5ml eppendorf at -40 °C, and this was further used as a sample for RNA isolation.

#### 2.2.2 RNA isolation, RT-PCR and gene copy estimation

RNA isolation from the pellet with the concentrated virus was performed using Nucleo-Spin® RNA Virus (Macherey-Nagel GmbH & Co. KG, Germany) isolation kit. MS2 phage was used as an internal control provided by TaqPathTM Covid-19 RT-PCR Kit. Some other specifics are, a) the nucleic acid was extracted by TaqPathTM Covid-19 RT-PCR Kit (Applied Biosystems), and Qubit 4 Fluorometer (Invitrogen) was used for RNA concentrations estimation, b) molecular process inhibition control (MPC) was evaluated through MS2 phage for the QC/QA analyses of nucleic acid extraction and PCR inhibition (Haramoto et al., 2018). We have described methodology elsewhere (Kumar et al., 2021 and 2020a). Briefly, steps were carried out as per the guideline provided with the product manual of Macherey-Nagel GmbH & Co. KG, and RNAs were detected using real-time PCR (RT-PCR).

SARS-CoV-2 gene was detected with Applied Biosystems 7500 Fast Dx Real-Time PCR Instrument (version 2.19 software) and for each run a template of 7 µl of extracted RNA was used with TaqPath™ 1 Step Multiplex Master Mix (Thermofischer Scientific, USA). Final reaction mixture (20 µmL) contained nuclease-free water 9 (10.50 µL), Master Mix (6.25 µL), and COVID-19 Real-Time PCR Assay Multiplex (1.25 µL). Positive control (TaqPath™ COVID 19 Control), negative control (from extraction run spiked with MS2), and no template control (NTC) were run with each batch. 40 cycles of amplification were set and results were interpreted based on the Ct values for three target genes i.e., ORF1ab, N Protein, and S Protein of SARS-CoV-2 along with that of MS2 used as an internal control.

Results were considered inconclusive if less than two genes are detected in the samples. Effective genome concentration was calculated semi-qualitatively using the equivalence of 500 copies of SARS-CoV-2 genes as 26 Ct-value (provided with the kit), and multiplying the RNA amount used as a template and the enrichment factor of waste water samples during the experimentation.

Test of significance and multivariate analyses (MVA) was performed with help of Statistical Package for the Social Sciences (SPSS 21) to evaluate the relatedness among various quality parameters analysed and to delineate the governing variables in the produced data-set. The OriginPro 2019b was used for data plotting and analysis.

## 3. Results and discussions

We detected and quantified variation in SARS-CoV-2 RNA from influent wastewater samples for two months (August and September) to understand the pandemic situation in Gandhinagar, Gujarat, India. Among 43 samples analyzed in the study, 40 were found positive, comprising two out of three target genes (**Table 1**). The distribution analysis of Ct values for different genes using boxplot is represented in **Fig.1a**. The average Ct values for S, N, and ORF 1ab genes were 32.66, 33.03, and 33.95, respectively. The Ct values of internal control (MS2 bacteriophage) ranged between 25.15 to 28.01. Also, no SARS-CoV-2 genes were detected in the negative control samples. The average gene copies were found to be maximum for S-gene (∼1223 copies/L), followed by N-gene (∼1022 copies/ L) and ORF 1 ab-gene (∼485 copies/L) (**Fig.1b**).

**Fig. 1.**
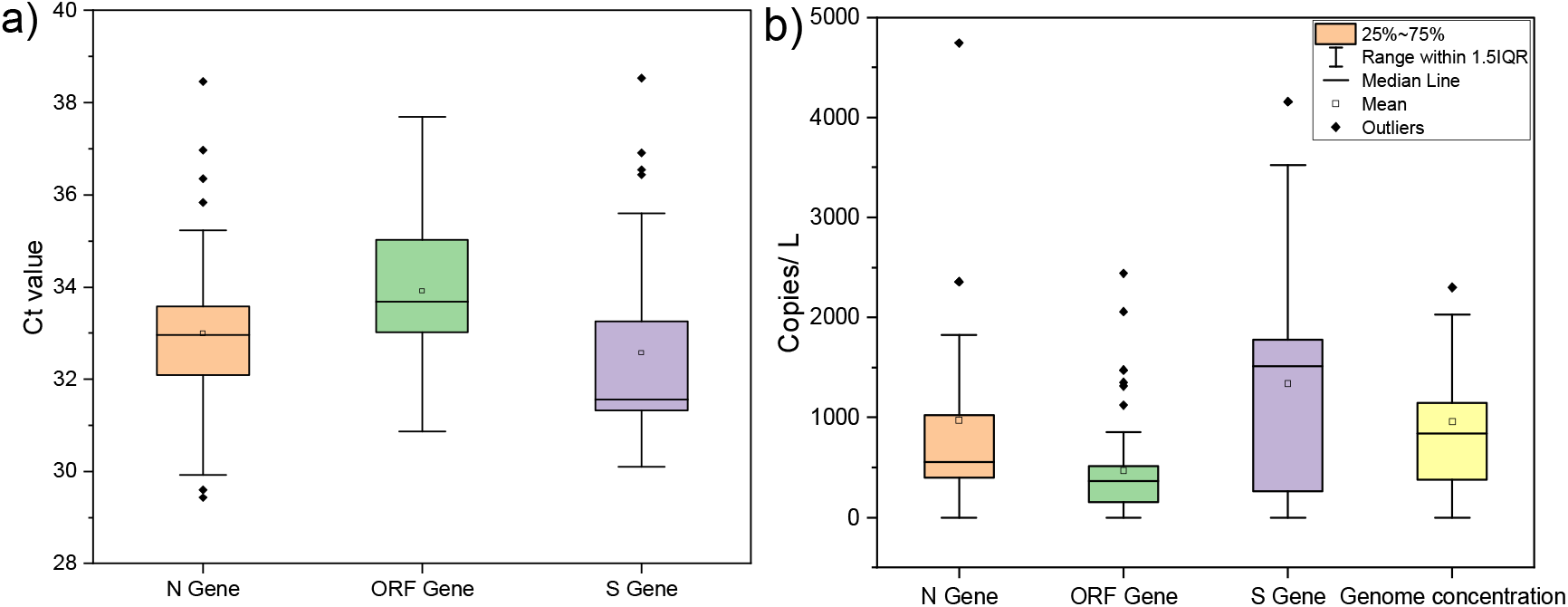
Distribution of SARS-CoV-2 viral gene a) Ct values, and b) target gene copies during entire study period in Gandhinagar.

**Table 1.**
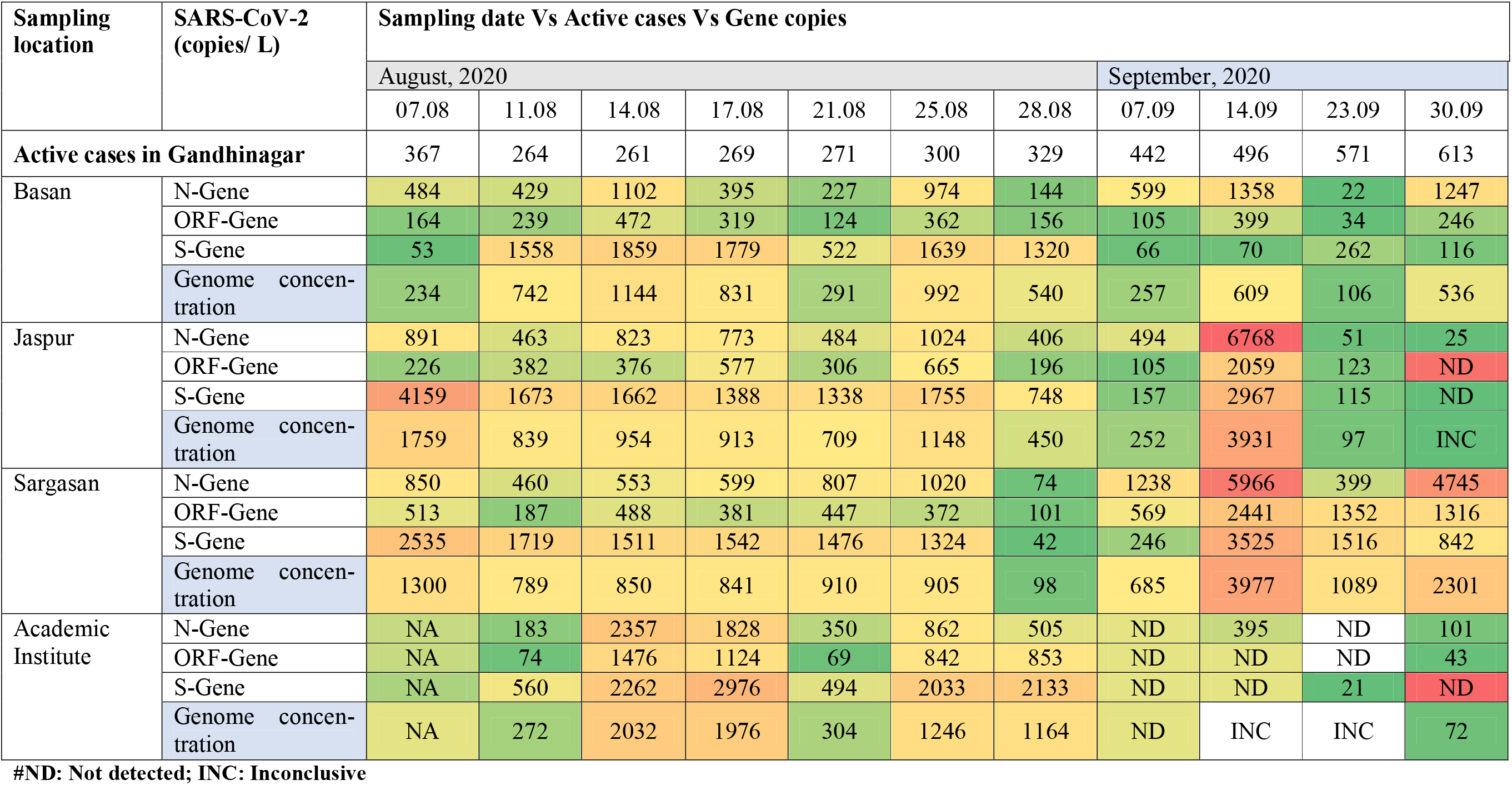
**Temporal variation in SARS-CoV-2 genetic material loading found in the influent and effluent samples collected from two different wastewater treatment plants**.

| Sampling location | SARS-CoV-2 (copies/ L) | Sampling date Vs Active cases Vs Gene copies |  |  |  |  |  |  |  |  |  |  |
| --- | --- | --- | --- | --- | --- | --- | --- | --- | --- | --- | --- | --- |
|  |  | August, 2020 |  |  |  |  |  |  | September, 2020 |  |  |  |
|  |  | 07.08 | 11.08 | 14.08 | 17.08 | 21.08 | 25.08 | 28.08 | 07.09 | 14.09 | 23.09 | 30.09 |
| Active cases in Gandhinagar |  | 367 | 264 | 261 | 269 | 271 | 300 | 329 | 442 | 496 | 571 | 613 |
| Basan | N-Gene | 484 | 429 | 1102 | 395 | 227 | 974 | 144 | 599 | 1358 | 22 | 1247 |
|  | ORF-Gene | 164 | 239 | 472 | 319 | 124 | 362 | 156 | 105 | 399 | 34 | 246 |
|  | S-Gene | 53 | 1558 | 1859 | 1779 | 522 | 1639 | 1320 | 66 | 70 | 262 | 116 |
|  | Genome concentration | 234 | 742 | 1144 | 831 | 291 | 992 | 540 | 257 | 609 | 106 | 536 |
| Jaspur | N-Gene | 891 | 463 | 823 | 773 | 484 | 1024 | 406 | 494 | 6768 | 51 | 25 |
|  | ORF-Gene | 226 | 382 | 376 | 577 | 306 | 665 | 196 | 105 | 2059 | 123 | ND |
|  | S-Gene | 4159 | 1673 | 1662 | 1388 | 1338 | 1755 | 748 | 157 | 2967 | 115 | ND |
|  | Genome concentration | 1759 | 839 | 954 | 913 | 709 | 1148 | 450 | 252 | 3931 | 97 | INC |
| Sargasan | N-Gene | 850 | 460 | 553 | 599 | 807 | 1020 | 74 | 1238 | 5966 | 399 | 4745 |
|  | ORF-Gene | 513 | 187 | 488 | 381 | 447 | 372 | 101 | 569 | 2441 | 1352 | 1316 |
|  | S-Gene | 2535 | 1719 | 1511 | 1542 | 1476 | 1324 | 42 | 246 | 3525 | 1516 | 842 |
|  | Genome concentration | 1300 | 789 | 850 | 841 | 910 | 905 | 98 | 685 | 3977 | 1089 | 2301 |
| Academic Institute | N-Gene | NA | 183 | 2357 | 1828 | 350 | 862 | 505 | ND | 395 | ND | 101 |
|  | ORF-Gene | NA | 74 | 1476 | 1124 | 69 | 842 | 853 | ND | ND | ND | 43 |
|  | S-Gene | NA | 560 | 2262 | 2976 | 494 | 2033 | 2133 | ND | ND | 21 | ND |
|  | Genome concentration | NA | 272 | 2032 | 1976 | 304 | 1246 | 1164 | ND | INC | INC | 72 |

### 3.1 The Monthly Variations

The monthly variation depicted a substantial rise in the average copies of N (∼120%) and ORF 1ab (∼38%) genes in the month of September as compared to August, while S-gene copies declined by 58% in September 2020 (**Fig.2 a**). The SARS-CoV-2 genome concentration was higher in the month of September (∼924.5 copies/ L) than August (∼897.5 copies/ L), corresponding to a ∼ 2.2-fold rise in the number of confirmed cases during the study period (**Fig. 2b**). Temporal variations in average SARS-CoV-2 gene copies were analyzed from different WWTPs in Gandhinagar are displayed in **Fig. 3a-d**. One-way ANOVA and Duncan post hoc test (**p < 0.05**) was performed to see the significance level in gene copy variation among different sampling dates. The results showed significant differences in N-gene (ANOVA, *F*= 2.68, p <0.05) and S-gene copies (ANOVA, *F*= 2.20, p <0.05) on the temporal scale (sampling dates). Conversely, differences were non-significant in case of ORF-1ab genes (ANOVA, *F*= 1.13, p >0.05) and genome concentration (ANOVA, *F*= 1.63, p >0.05).

**Fig. 2.**
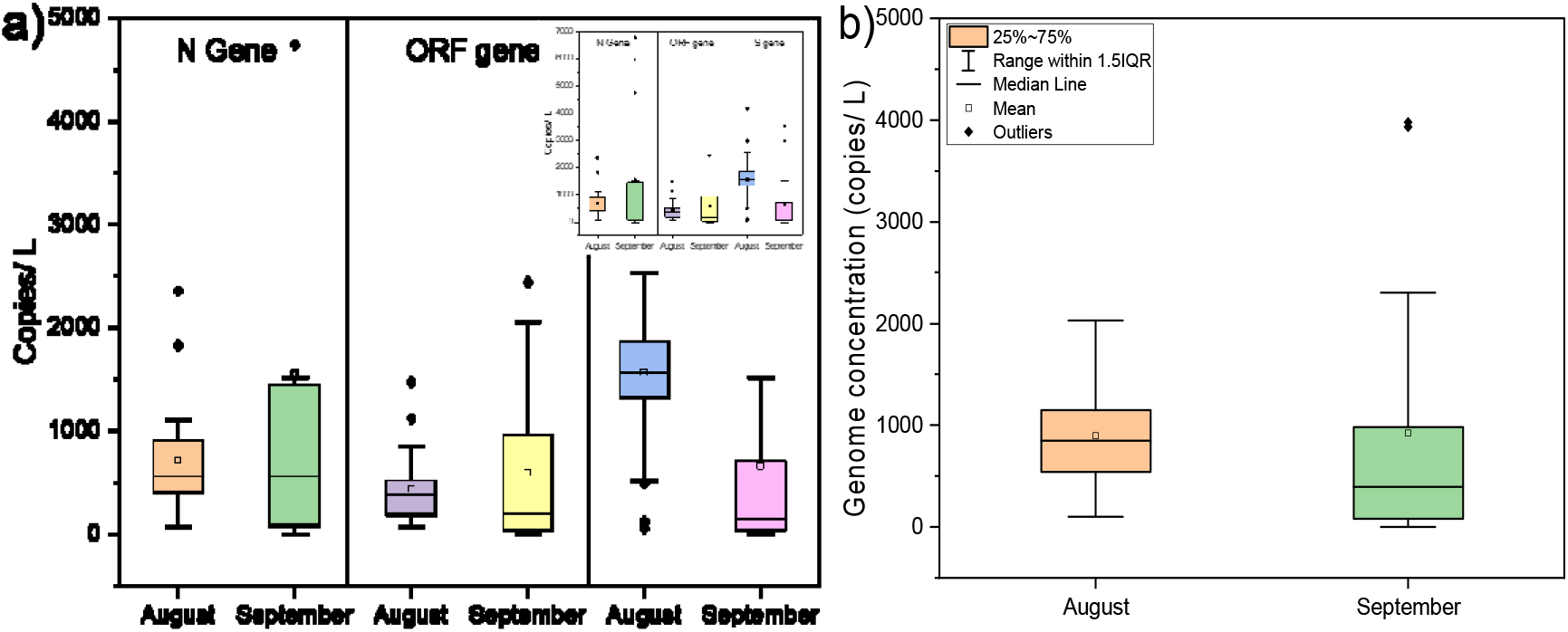
Distribution of SARS-CoV-2 gene copies on a temporal scale (monthly variation), a) Target gene copies, b) Genome concentration.

**Fig. 3.**
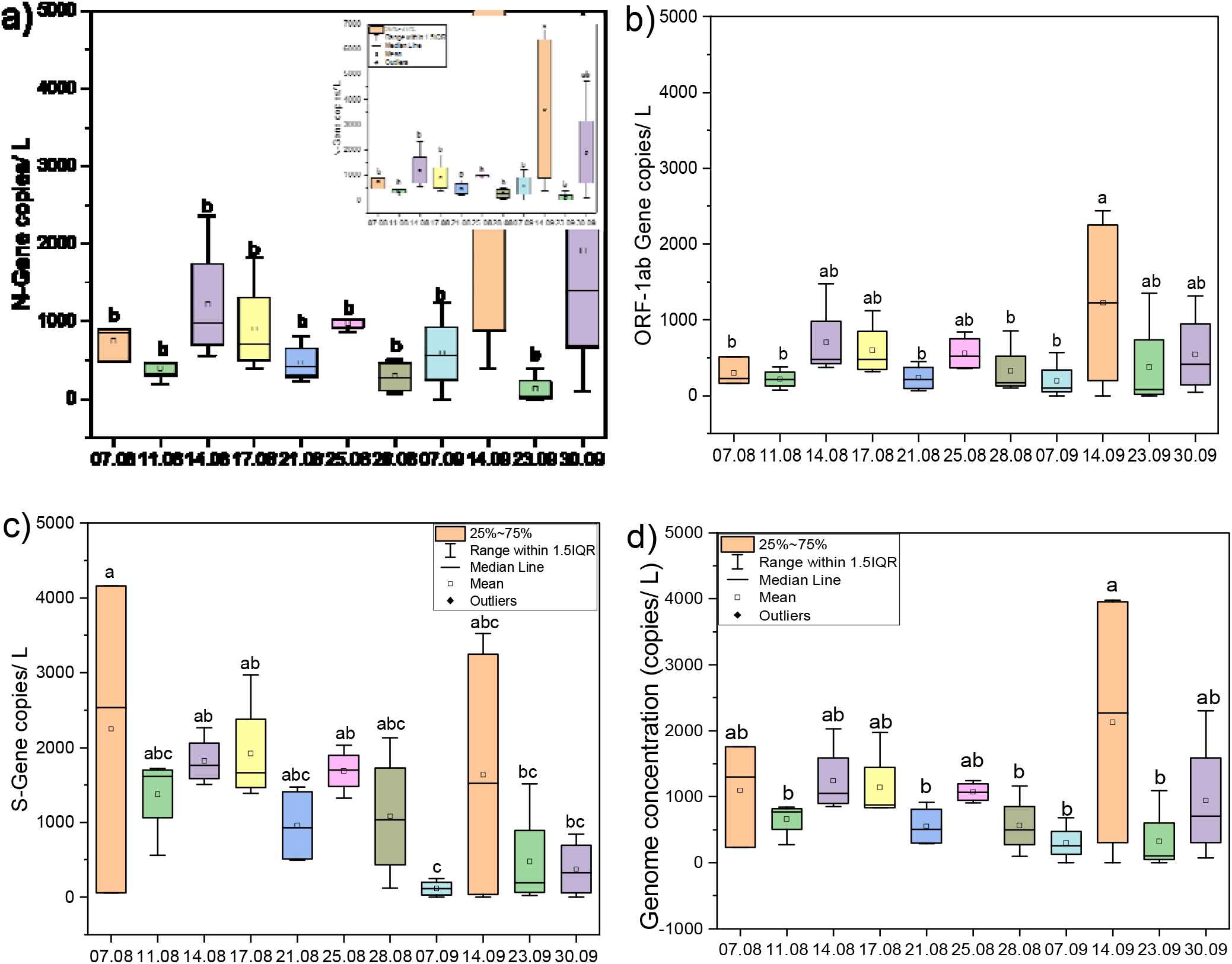
Temporal variations in average SARS-CoV-2 gene copies collected from different STPs in Gandhinagar, a) N-Gene, b) ORF1 ab Gene, c) S-Gene, and d) Genome concentration. Alphabetical letters in graphs represent a statistically significant difference of **p < 0.05** by Duncan post hoc test

There are some studies available around the globe on early detection of SARS-CoV-2 RNA in wastewater, even before the first report of clinical diagnosis. For example, Madema et al. (2020) reported the presence of SARS-CoV-2 genetic material in wastewater in February, even before the official declaration of the first case in the Netherlands. Likewise, La Rosa et al. (2020) reported SARS-CoV-2 genetic material in wastewater samples before the first official documented report from two different cities in Italy. Similarly, Randazzo et al. (2020) detected SARS-CoV-2 RNA in wastewater samples from Spain. Since then, many researchers detected and reported the occurrence of SARS-CoV-2 RNA in wastewater samples and pondered its applicability for WBE surveillance (Ahmed et al., 2020, Kumar et al. 2020a,b). However, a few studies available focused on assessing its potential on the temporal scale in relation to the changes in COVID cases.

### 3.2 The Early Warning Capability

In this view, the present research work followed our first proof of the concept, where we detected SARS-CoV-2 genetic material in wastewater and proposed its wide applicability for COVID surveillance in the community (Kumar et al. 2020a). Examining the potential of WBE for COVID-19 surveillance as a potential tool showed that the percentage change in genome concentration level on a particular date was in conjunction with the confirmed cases registered 1-2 weeks later on a temporal scale by the regulatory authority based on clinical tests (**Fig. 4**). For example, on August, 21^st^, 2020, a sharp decline of ∼51% was noticed in the percentage change in the average genome concentration which was followed by ∼0.76% decline in the percentage change in confirmed COVID cases on August, 28^th^, 2020. Likewise, on August 25^th^, 2020, a steep hike of ∼75% in the percentage change in the average genome concentration was noticed, which was followed by ∼11% increment in the percentage change in confirmed COVID cases on September 7^th^, 2020. Therefore, we can predict the severity of the pandemic situation 1-2 weeks prior to the official reports by the regulatory body based on clinical tests. The results unravel the potential of WBE surveillance of COVID-19 as an early warning tool displayed by the adequate presence of SARS-CoV-2 genetic material in wastewater samples though limited cases were documented and based on the immediate future trends. These findings were in agreement with those of Ahmed et al. (2020b), who noticed a longitudinal decline in the presence of SARS-CoV-2 RNA with the tapering of the first epidemic wave; however, there was no concrete relationship between virus RNA and daily cases numbers.

**Fig. 4.**
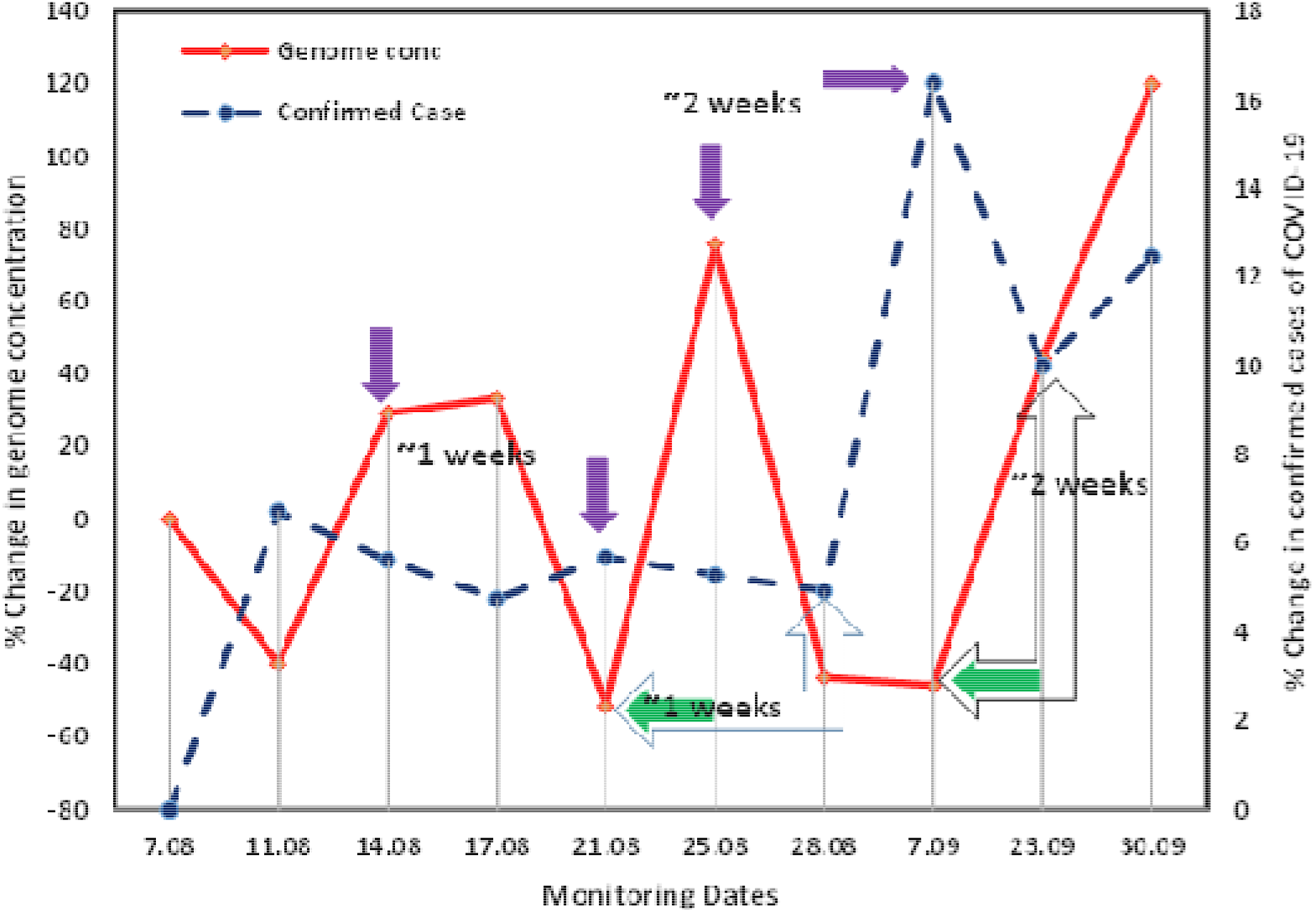
Potential and evidence of wastewater-based epidemiology surveillance of Covid-19 pandemic as an early warning tool in Gandhinagar

### 3.1 Relatedness with COVID-19 cases and water quality through multivariate analyses

Finally, MVA was performed to know the relation among influent wastewater physicochemical characteristics, SARS-CoV genetic material, and pandemic status (i.e., confirmed, active, recovered, and deceased cases) through principal component analysis depicted by PCs loading in a 3-D domain during the entire two months of the monitoring period **(Fig. 5a and b)**. A summary description of in-situ parameters **(Table S3)**, variation explained, eigenvalue variations, and principal component loadings for influent wastewater **(Table S4, Fig S1)** have been provided as supplementary items.

**Fig. 5.**
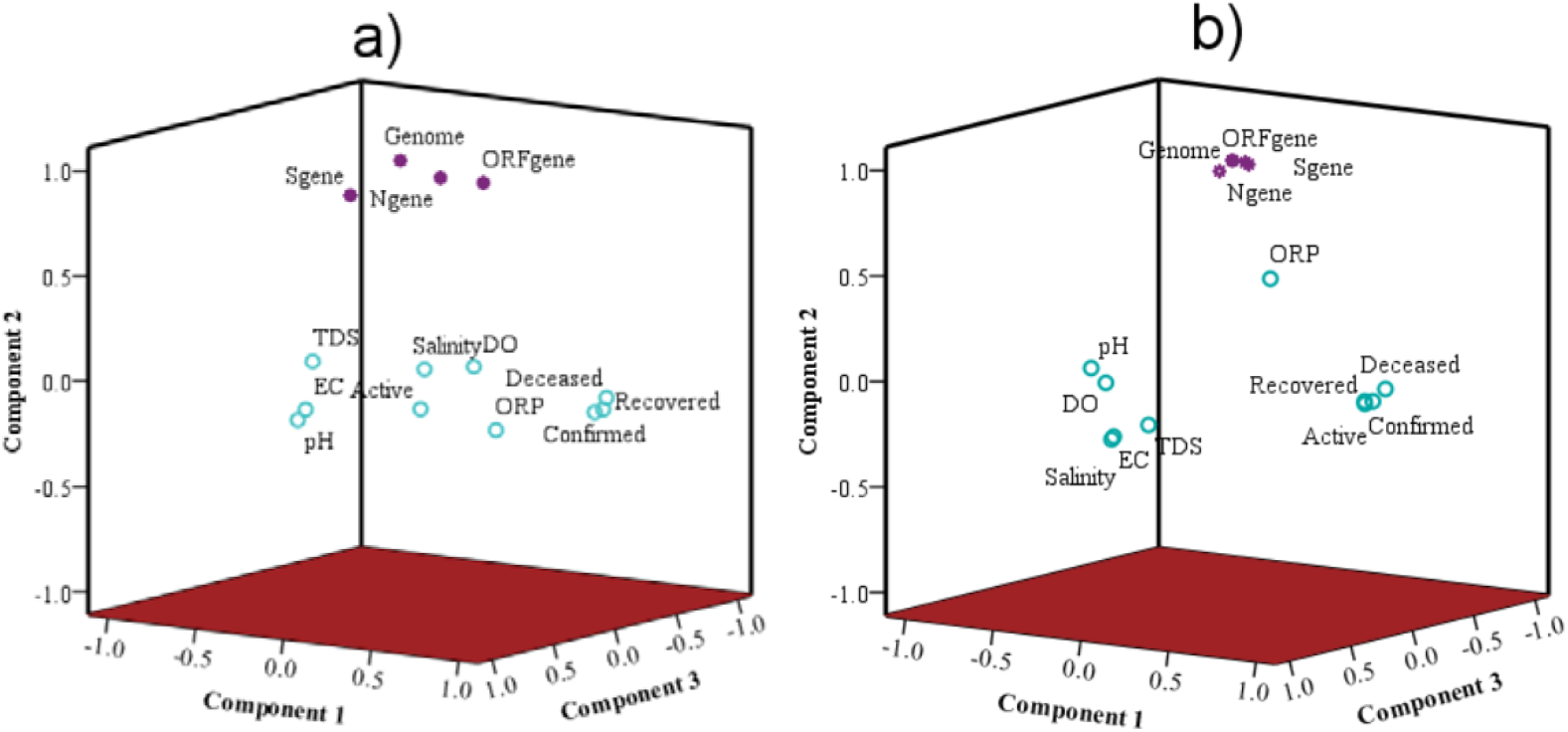
Principal component analysis (PCA) to show relation/ interaction among physicochemical parameters of untreated wastewater and targeted SARS-CoV-2 gene copies, a) August; b) September

Principal component analyses show a comprehensive picture of the overall interaction among SARS-CoV-2 genetic material and influent wastewater characteristics. The entire dataset obtained for August and September were subjected to PCA and projected in the 3-D domain of three main PCs. In the month of August, four PCs were identified that explains 76.9% of the total variance in the dataset. The first PCs explained 26.9% of the total variance with significant loading for COVID cases forming a cluster (confirmed, recovered, and active cases) with moderate loading (∼0.5) of influent wastewater parameters (ORP and DO) and weak loading for ORF gene (**Fig. 5a** and Supplementary **Table S3** and **S4**). On the other hand, nearly the same (∼23.8%) variation of data sets is explained by SARS-CoV-2 genes, and genome concentrations form a cluster upper left domain with significant loadings for effective genome concentrations (0.98) followed by ORF-1ab, N-genes, and S-genes as PC2. Interestingly in August, the ORF 1ab genes illustrated positive loadings in both PC1 and PC2. The PC3 and PC4 exhibited almost similar contribution (∼13%) of the total variance.

In September, the complexion changed significantly with the overall reductions of PCs to three, explaining cumulative variations of 84% in the dataset. The trends were almost similar to the month of August. However, SARS-CoV-2 genes exhibited higher loadings. Order of loadings among SARS-CoV-2 genes and genome remains same i.e., effective genome concentration> ORF-1ab> N-genes> S-genes. The confirmed and active COVID cases showed a positive relationship with SARS-CoV-2 genes (ORF 1ab, N-gene, and genome concentration), though the relationship was not strong due to weak correlation coefficients (<0.1). Though the results of MVA suggested a weak relationship between the SARS-CoV-2 genome concentration and confirmed cases, yet the percentage change in genome concentration showed a positive relation to the percentage change in the confirmed cases. This might be because of the effect of change in SARS-CoV-2 genome concentration was reflected in 1 to 2 weeks later data of the confirmed cases. This could also partly be ascribed to the number of confirmed cases not necessarily reflect the actual prevalence of the disease (Hata et al. 2020).

## 4. Conclusion

A temporal variation of SARS-CoV-2 RNA presence in influent wastewater was studied for a period of two months in Gandhinagar, India. Out of 43 samples, 40 samples were found positive, while RT-PCR showed greater sensitivity for S-gene, followed by N-gene and ORF 1ab gene. A comparison of monthly variation demonstrated higher SARS-CoV-2 genome concentration in September (∼924.5 copies/ L) than August (∼897.5 copies/ L) in line with the ∼2.2-fold rise in the number of confirmed cases during the study period. The results profoundly unravel the potential of WBE surveillance to predict the fluctuation of COVID-19 cases to provide an early warning. Our study explicitly suggests that it is the need of hour that the wastewater surveillance must be included as an integral part of COVID-19 pandemic monitoring which can not only help the water authorities to identify the hotspots within a city but can provide up to 2 weeks of time lead for better tuning the management interventions.

## Notes

The authors declare no competing financial interest.

## Data Availability

Although the paper contains all the basic data, we will provide it more details on correspondence.

## Acknowledgement

This work is funded by Kiran C Patel Centre for Sustainable Development at IIT Gandhinagar, UNICEF, Gujarat and UK-India Education and Research Initiative (UK-IERI). We acknowledge the help received from Dr. Vaibhav Srivastava who contributed towards sample and data analyses.

## References

Ahmed, W., Angel, N., Edson, J., Bibby, K., Bivins, A., O’Brien, J.W., Choi, P.M., Kitajima, M., Simpson, S.L., Li, J. and Tscharke, B., 2020a. First confirmed detection of SARS-CoV-2 in untreated wastewater in Australia: A proof of concept for the wastewater surveillance of COVID-19 in the community. Science of The Total Environment, p.138764. https://doi.org/10.1016/j.scitotenv.2020.138764

Ahmed, W., Tscharke, B., Bertsch, P.M., Bibby, K., Bivins, A., Choi, P., Clarke, L., Dwyer, J., Edson, J., Nguyen, T.M.H. and O’Brien, J.W., 2020b. SARS-CoV-2 RNA monitoring in wastewater as an early warning system for COVID-19 transmission in the community: a temporal case study. Science of The Total Environment, p.144216. https://doi.org/10.1016/j.scitotenv.2020.144216

Chan, P.K., Lui, G., Hachim, A., Ko, R.L., Boon, S.S., Li, T., Kavian, N., Luk, F., Chen, Z., Yau, E.M. and Chan, K.H., 2020. Serologic responses in healthy adult with SARS-CoV-2 reinfection, Hong Kong, August 2020. Emerging infectious diseases, 26(12), p.3076. DOI: 10.3201/eid2612.203833

Cheung, K.S., Hung, I.F., Chan, P.P., Lung, K.C., Tso, E., Liu, R., Ng, Y.Y., Chu, M.Y., Chung, T.W., Tam, A.R. and Yip, C.C., 2020. Gastrointestinal manifestations of SARS-CoV-2 infection and virus load in fecal samples from the Hong Kong cohort and systematic review and meta-analysis. Gastroenterology. https://doi.org/10.1053/j.gastro.2020.03.065

Gupta, M.K., Vemula, S., Donde, R., Gouda, G., Behera, L. and Vadde, R., 2020. In-silico approaches to detect inhibitors of the human severe acute respiratory syndrome coronavirus envelope protein ion channel. Journal of Biomolecular Structure and Dynamics, pp.1–11. https://doi.org/10.1080/07391102.2020.1751300

Haramoto, E., Malla, B., Thakali, O., Kitajima, M., 2020. First environmental surveillance for the presence of SARS-CoV-2 RNA in wastewater and river water in Japan. Sci. Total. Environ. 737, 140405. https://doi.org/10.1016/j.scitotenv.2020.140405

Hata, A., Hara-Yamamura, H., Meuchi, Y., Imai, S. and Honda, R., 2020. Detection of SARS-CoV-2 in wastewater in Japan during a COVID-19 outbreak. Science of The Total Environment, p.143578. https://doi.org/10.1016/j.scitotenv.2020.143578

Havers, F.P., Reed, C., Lim, T., Montgomery, J.M., Klena, J.D., Hall, A.J., Fry, A.M., Cannon, D.L., Chiang, C.F., Gibbons, A. and Krapiunaya, I., 2020. Seroprevalence of antibodies to SARS-CoV-2 in 10 sites in the United States, March 23-May 12, 2020. JAMA, 180(12), pp.1576–1586. DOI:10.1001/jamainternmed.2020.4130

Kumar, M., Patel, A.K., Shah, A.V., Raval, J., Rajpara, N., Joshi, M. and Joshi, C.G., 2020a. First proof of the capability of wastewater surveillance for COVID-19 in India through detection of genetic material of SARS-CoV-2. Science of The Total Environment, 746, p.141326. https://doi.org/10.1016/j.scitotenv.2020.141326

Kumar, M., Thakur, A.K., Mazumder, P., Kuroda, K., Mohapatra, S., Rinklebe, J., Ramanathan, A.L., Cetecioglu, Z., Jain, S., Tyagi, V.K. and Gikas, P., 2020b. Frontier review on the propensity and repercussion of SARS-CoV-2 migration to aquatic environment. Journal of Hazardous Materials Letters, 1, p.100001. https://doi.org/10.1016/j.hazl.2020.100001

Kumar, M., Kuroda, K., Patel, A.K., Patel, N., Bhattacharya, P., Joshi, M. and Joshi, C.G., 2020. Decay of SARS-CoV-2 RNA along the wastewater treatment outfitted with Upflow Anaerobic Sludge Blanket (UASB) system evaluated through two sample concentration techniques. Science of the Total Environment, 754, p.142329. https://doi.org/10.1016/j.scitotenv.2020.142329

La Rosa, G., Iaconelli, M., Mancini, P., Ferraro, G.B., Veneri, C., Bonadonna, L., Lucentini, L. and Suffredini, E., 2020. First detection of SARS-CoV-2 in untreated wastewaters in Italy. Science of The Total Environment, p.139652. https://doi.org/10.1016/j.scitotenv.2020.139652

Lavezzo, E., Franchin, E., Ciavarella, C., Cuomo-Dannenburg, G., Barzon, L., Del Vecchio, C., Rossi, L., Manganelli, R., Loregian, A., Navarin, N. and Abate, D., 2020. Suppression of a SARS-CoV-2 outbreak in the Italian municipality of Vo’. Nature, 584(7821), pp.425–429. https://doi.org/10.1038/s41586-020-2488-1

Medema, G., Heijnen, L., Elsinga, G., Italiaander, R., Brouwer, A., 2020. Presence of SARS Coronavirus-2 RNA in sewage and correlation with reported COVID-19 prevalence in the early stage of the epidemic in the Netherlands. Environ Sci Technol Lett. 7 (7), 511–516. https://doi.org/10.1021/acs.estlett.0c00357

Mizumoto, K. and Chowell, G., 2020. Estimating Risk for Death from Coronavirus Disease, China, January–February 2020. Emerging infectious diseases, 26(6), p.1251. DOI: 10.3201/eid2606.200233

Ni, W., Yang, X., Yang, D., Bao, J., Li, R., Xiao, Y., Hou, C., Wang, H., Liu, J., Yang, D. and Xu, Y., 2020. Role of angiotensin-converting enzyme 2 (ACE2) in COVID-19. Critical Care, 24(1), pp.1–10. https://doi.org/10.1186/s13054-020-03120-0

Nishiura, H., Linton, N.M. and Akhmetzhanov, A.R., 2020. Serial interval of novel coronavirus (COVID-19) infections. International journal of infectious diseases. https://doi.org/10.1016/j.ijid.2020.02.060

Randazzo, W., Truchado, P., Cuevas-Ferrando, E., Simón, P., Allende, A., Sánchez, G., 2020a. SARS-CoV-2 RNA in wastewater anticipated COVID-19 occurrence in a Low prevalence area. Water Res. 181, 115942. https://doi.org/10.1016/j.watres.2020.115942

Rimoldi, S. G., Stefani, F., Gigantiello, A., Polesello, S., Comandatore, F., Mileto, D., Maresca, M., Longobardi, C., Mancon, A., Romeri, F., Pagani, C., Moja, L., Gismondo, M.R., Salerno, F.,2020. Presence and vitality of SARS-CoV-2 virus in wastewaters and rivers. Science of the Total Environment, 744:140911

Wurtzer, S., Marechal, V., Mouchel, J.M., Maday, Y., Teyssou, R., Richard, E., Almayrac, J.L. and Moulin, L., 2020. Evaluation of lockdown impact on SARS-CoV-2 dynamics through viral genome quantification in Paris wastewaters. medRxiv. https://doi.org/10.1101/2020.04.12.20062679

Wong, M.C., Huang, J., Lai, C., Ng, R., Chan, F.K. and Chan, P.K., 2020. Detection of SARS-CoV-2 RNA in fecal specimens of patients with confirmed COVID-19: a meta-analysis. Journal of Infection. https://doi.org/10.1016/j.jinf.2020.06.012

World Health Organization Novel Coronavirus (2019-nCoV) Situation Report – 1 (2020) https://www.who.int/emergencies/diseases/novel-coronavirus-2019/situation432-reports

Xiao, F., Tang, M., Zheng, X., Liu, Y., Li, X. and Shan, H., 2020. Evidence for gastrointestinal infection of SARS-CoV-2. Gastroenterology, 158(6), pp 1831–1833. https://doi.org/10.1053/j.gastro.2020.02.055

Yang, R., Gui, X. and Xiong, Y., 2020. Comparison of clinical characteristics of patients with asymptomatic vs symptomatic coronavirus disease 2019 in Wuhan, China. JAMA Network Open, 3(5), pp.e2010182–e2010182. DOI:10.1001/jamanetworkopen.2020.10182

Zhang, N., Gong, Y., Meng, F., Bi, Y., Yang, P. and Wang, F., 2020. Virus shedding patterns in naso- pharyngeal and fecal specimens of COVID-19 tients. MedRxiv. https://doi.org/10.1101/2020.03.28.20043059

